# Evaluating the effects of supervised consumption sites on housing prices in Montreal, Canada using controlled interrupted time series and hedonic price models

**DOI:** 10.1101/2023.06.09.23291209

**Authors:** Maximilian Schaefer, Dimitra Panagiotoglou

**Affiliations:** Department of Epidemiology, Biostatistics, and Occupational Health, McGill University, Montreal, Canada

**Keywords:** Harm reduction, opioid use, opioid overdose prevention, real estate value, spatio-temporal analysis

## Abstract

**Background:** In 2017, three brick and mortar supervised consumption sites (SCS) began operating in Montreal, Canada. Opponents argued the sites would attract people who use drugs to the respective neighbourhoods, contributing to reductions in residential real estate values.

**Methods:** We used controlled interrupted time series and hedonic price models to evaluate the effects of Montreal’s SCSs on residential real estate. We linked the Quebec Professional Association of Real Estate Brokers’ housing sales data provided by Centris Inc. with Statistics Canada’s census tract data, neighbourhood proximity measures, and Canadian Urban Environmental Health Research Consortium’s gentrification measures. We restricted analysis to sales between 1 January 2014 and 31 December 2021, and within 1,000m of a SCS (treated) or a men’s homeless shelter (control). We controlled for internal (e.g., number of bed/bathrooms, unit size) and external attributes (e.g., neighbourhood demographics; proximity to amenities), and included a spatio-temporal lag to account for correlation between sales.

**Results:** When controlling for census tract data and gentrification measures, the price of homes sold immediately after SCSs were implemented was 5.2% lower (95% CI: -1.4%, -8.8%) compared with control sales (level effect). However, the monthly value increased 0.6% faster (95% CI: 0.4%, 0.7%) in treated neighbourhoods (trend effect). Compared with the counterfactual (i.e., SCS never implemented), sales in treated neighbourhoods observed an absolute increase of $37,931.86 (95% CI: $12,223.35, $138,088.50) by December 2021. When we also controlled for proximity scores, the immediate level effect post-implementation disappeared (−3.3%, 95% CI: -0.7%, 1.1%), but monthly trend gains persisted (0.9%, 95% CI: 0.7%, 1.0%).

**Conclusion:** We observed a modest negative effect on prices immediately following SCS implementation. However, controlling for proximity to neighbourhood amenities eliminated the level effect. Positive month-on-month price gains were consistently observed, suggesting community wide benefits of SCS implementation.

## Background

In 2017, Montreal, Canada, introduced three brick and mortar supervised consumption sites (SCSs) as part of its harm reduction program with the aim to reduce the negative effects of illicit drug use (Strike & Watson, 2019). Federally approved and staffed by medical professionals trained in addictions medicine, SCSs provide critical overdose reversal services, overdose education and naloxone distribution, clean drug use equipment and disposal of used items, primary care services, safe injection practices and wound care education, and housing and employment support (Government of Canada, 2018).

Despite extensive evidence demonstrating the benefits of SCSs on the health and well-being of people who use drugs (PWUD), SCSs remain politically controversial (Potier C et al., 2014). Local politicians, residents and business owners vehemently resist their implementation (Cruz MF et al., 2007; Small D, 2007). Opponents believe SCSs attract PWUD to the sites’ neighbourhoods (the ‘honey-pot effect’) and argue this influx of PWUD increases local crime, contributes to physical and aesthetic deterioration, and reduces property value (Kolla et al., 2017; Williams & Ouellet, 2010). Proponents of SCSs try to alleviate concerns of the honey-pot effect citing evidence that SCS clients typically reside within 500m of the site (Marshall BDL et al., 2011; The Evaluation of Overdose Prevention Sites Working Group & Lori Wagar, 2018) and stress sites are situated in high-risk neighbourhoods with a known PWUD presence (Supervised Consumption Services Review Committee & Alberta Health, 2020). Supporters also refer to the handful of studies that show no changes in property crime, marginal increases in small-scale drug-dealing, and reductions in public drug use after SCS implementation (Kennedy et al., 2017; Kimber et al., 2005).

Less easily addressed are opponents’ skepticism on the generalizability of results, given most studies have focused on the effects of harm reduction interventions in Vancouver’s Downtown Eastside (home to Canada’s only SCS prior to 2016), and researchers’ disregard for ‘the interests of larger communit[ies]’ (Kolla et al., 2017). Even where community stakeholders acknowledge that these facilities have positive health effects, ‘not in my backyard’ (NIMBY) resistance persists. Opponents fixate on SCSs’ potential to tarnish their communities’ reputations more than public drug use; and maintain the downstream consequences are reduced small-business patronage and home values. Given NIMBY sentiments stem from complex social, cultural and political perspectives, it is important to understand the effects of harm reduction interventions on local communities (Bosque-Prous & Brugal, 2016). With minimal exploration of the effects of SCSs on neighbourhoods; (Liang & Alexeev, 2023) these perceived threats have repeatedly barred agencies from operating SCSs in high-risk communities (Guye A, 2021; Supervised Consumption Services Review Committee & Alberta Health, 2020).

Considering the ongoing tensions, we tested the hypothesis that SCSs have a negative effect on residential real estate value in Montreal using the city’s recently implemented SCSs as a natural experiment.

## Methods

Montreal is an ideal setting to study the effects of SCSs on residential real estate. The city has one of the largest and most complex PWUD populations in Canada. Approximately 4,000 people inject drugs (Leclerc P et al., 2014), the proportion of people who inject daily remains high, and roughly 50% of PWUD consume drugs other than opioids (Bruneau J et al., 2012). Further, the PWUD population is geographically scattered (Green T, 2003), a necessary condition for the purported honey-pot effect. Finally, unlike Vancouver and Toronto’s housing markets which were exceptionally hot in recent years, Montreal’s market has enjoyed steady but modest gains, thus making it more representative of other Canadian cities and less likely to obfuscate the effects of SCSs on residential real estate value.

We used a controlled interrupted times series study design for this study. We included residential real estate sales records from 1 January 2014 to 31 December 2021, to allow sufficient time to observe the effects of SCSs on prices post-implementation, while limiting the potential for large changes in neighbourhood demographics.

We designated sales that occurred within 1,000m of SCSs’ locations as treated units, and sales that occurred within 1,000m of men’s homeless shelters in operation since 2010 as control units. We selected men’s homeless shelters as control sites because there is considerable demographic overlap (e.g., age, employment status, mental health needs) between SCSs’ and men’s homeless shelter clients; many SCSs are based in shelters, and similar NIMBY resistance dominates discussions of new homeless shelter sites (Oakley, 2017). Although we also considered needle and syringe program sites for our controls, the greater variation in where services are offered (e.g., hospitals, pharmacies, health clinics, public health units, community based organizations, and secondary distributors) combined with differences in how they are used (e.g., frequency and duration of visits)(Klein A, 2007) made them less suitable controls.

Residential real estate sales data were provided by Centris Inc. which captures over 89.6% of all residential real estate sales for the island of Montreal. Records included each home’s listed and purchased price, the duration on the market, number of bedrooms and bathrooms, total living space and property size, year of build, and noteworthy features (e.g., number of parking spots, separate garage, historical building). We excluded sales within 1,000m of both treated and control catchments to minimize spillover effects in our analysis (Figure 1). As this database includes sales that were not at ‘arm’s length’ (e.g., property with a sale price of CAD $1), we removed outliers whose final sale price was less than $20,000. We also excluded units with no listed total living space, fewer than two or more than twenty rooms, where number of bathrooms was missing or zero, number of bedrooms was missing, number of bathrooms or bedrooms exceeded the total number of rooms listed, and sales with missing civic numbers or street names.

**Figure 1:**
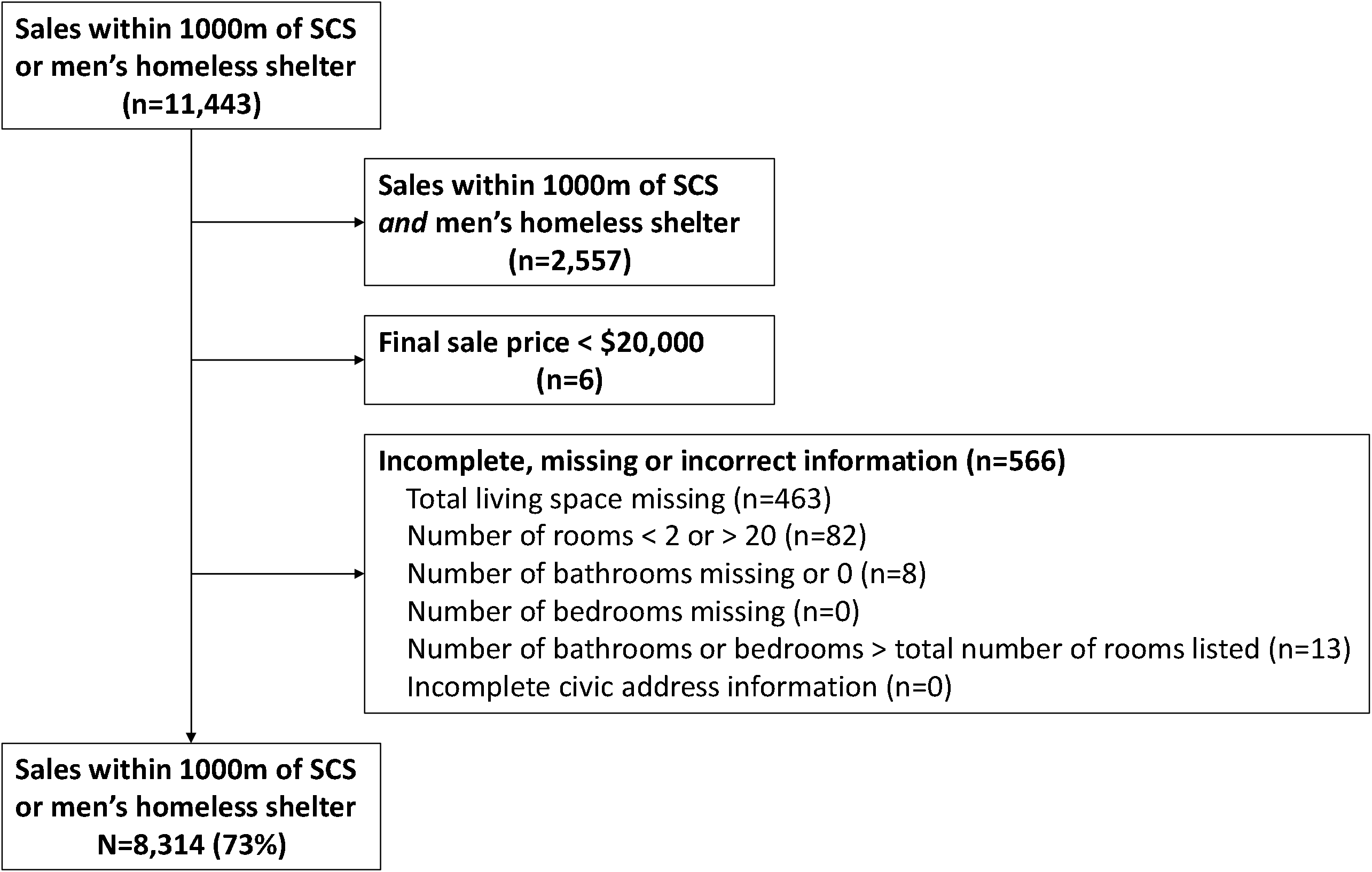
Sales record inclusion flowchart

We linked the sales records with Statistics Canada’s 2016 census tract data summarizing neighbourhoods’ demographics (i.e., median age of population, average household size, median total income, and proportion of the population that are visible minorities, did not complete secondary school, have a post-secondary education, and are unemployed)(Statistics Canada, 2023) and the Canadian Housing and Mortgage Corporation’s proximity measures database which provide normalized scores for distance to amenities (i.e., primary schools, childcare, grocery stores, healthcare, pharmacies, public parks, and public transit)(Statistics Canada & Canada Mortgage and Housing Corporation, 2020). The proximity scores are based on distance to services, and size of services using a simple gravity model. The resulting normalized index value for each amenity is on a scale of 0 to 1, with 1 demonstrating the highest proximity in Canada (Statistics Canada & Canada Mortgage and Housing Corporation, 2020). Finally, we included the Canadian Urban Environmental Health Research Consortium’s gentrification measures which identify areas at risk of or which have recently undergone gentrification within Canada(Firth et al., 2020).

Because sales prices were not normally distributed, we used the semi-log functional form for our models. This form allows the logarithm to be made for either the dependent or independent variables and enables simple interpretation of outputs.

There is a substantial body of literature that uses hedonic price models to identify both internal and external attributes that affect housing prices. Using these models, studies demonstrate buyers will pay a premium for proximity to desired amenities (e.g., schools, commercial centres, revived city centres) (Ding X et al., 2020; Dubé et al., 2017) and sellers will incur a penalty for proximity to dis-amenities (e.g., airports, homeless shelters) (Batóg et al., 2019; Galster et al., 2004). The relationship between amenities and housing prices is so robust that *announcements* of future amenities impact housing prices (Mense A & Kholodilin, 2014; Yen et al., 2018).

In our regression model the β estimates represent percentage differences in the closing price, not dollar amounts:

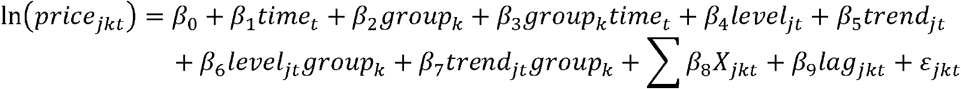

Where *β*_0_ is the semi-log price intercept for control sales; *β*_1_ is the pre-intervention time trend *t* observed for control sales; *β*_2_ is the pre-intervention difference between treated and control sales’ intercepts where *k*=0 for controls, and *k*=1 for treated units; *β*_3_ is the pre-intervention difference between treated and control sales trends; *β*_4_ and *β*_5_ describe the level and trend changes in the control group post-intervention implementation relative to pre-intervention, respectively; *β*_6_ is the level difference in sales price for treated units relative to the observed price change for the control units; *β*_7_ is the difference in time trend for treated sales relative to the difference in control price trends observed post-intervention; ∑ *β*_8_*X*_*jkt*_ is a vector of internal and external housing attributes (e.g., floor size, number of bed/bathrooms, distance to SCS or shelter, and neighbourhood proximity scores); *β*_9_ is the spatial-temporal price lag; and *ε*_*jkt*_ is the residual error term.

To create the spatio-temporal price lag for each sale we used the methods proposed by Higgins et al., 2019 (Higgins et al., 2019). Briefly, we calculated the spatial proximity between each combination of sales [i,j] using each sale’s geo-coordinates and the Euclidean distance method. Based on results from the Global Moran’s I test, we determined the spatial effects of neighbouring transaction *j* on closing price of sale *i* became negligible beyond 1,000m and set this as our cut-off. Combinations of sales [i,j] in the same complex were given a correlation value of 1, sales within the cut-off were assigned the inverse of their distance, and sales beyond the 1,000m threshold for correlation were assigned a value of 0 influence. We then pooled the values between each sale combination’s spatial weight into matrix *S*.

To account for temporal associations between sales [i,j] we applied Dube and Legros’ 2013 method (Dubé & Legros, 2013). We sorted all the sales in chronological order and allowed for temporal associations between transactions *i* and *j* that occurred up to 12 months in the past, and six months in the future. Doing so compensates for plausible ‘anchoring’ behaviour where property owners set their asking price not just on prices secured in comparable sales in the past, but on future expectations as well (Higgins et al., 2019). We pooled each sale combination’s temporal weight into matrix *T*. Then using the Hadamard product on matrices *S*⊙*T*, we created the spatio-temporal weight matrix *W*. We normalized *W* using spectral transformation for row-standardization (Higgins et al., 2019) before applying the weight to a matrix of all closing prices to create each sales record’s price lag.

Distances between sales records and treatment and control locations were confirmed using geocoding and distance matrices in QGIS. Addresses were geocoded to retrieve geographic coordinates using Nominatim/Open Street Maps. Coordinates were then used to create distance matrices between treatment and control locations, and sales records. Data were organized and models were run in R (version 4.2.2) within R Studio (version 2023.03.0) using tidyverse, readxl, openxlsx, tableone, olsrr, car, ape, spdep, geoR, lubridate, leaps, twang, scales, readr, data.table, and flextable packages.

We ran our models under two sets of conditions due to missing data in the proximity measures database. In the first set, we included all records that successfully linked with neighbourhood census tract data and gentrification measures (n=6,903). In the second set, we included only records that also successfully linked with proximity scores (n=4,815). We performed all analyses for both sets of models.

This study was exempt from ethics review by McGill University’s Institutional Review Board.

## Results

Our analysis included 2,209 treated sales and 4,694 control sales in the first set of models, and 2,057 (−6.9%) treated sales and 2,758 (−41.2%) control sales in the subset of models. Of records in the first set, 207 (3.0%) met our definition for house flipping – homes purchased and resold within two years: 165 of these were flipped once, and 42 were flipped twice, i.e., sold three times within two years each. The mean closing price pre-intervention was $271,473.73 and $419.001.00 in the treated and control catchments, respectively (Table 1).

**Table 1:**
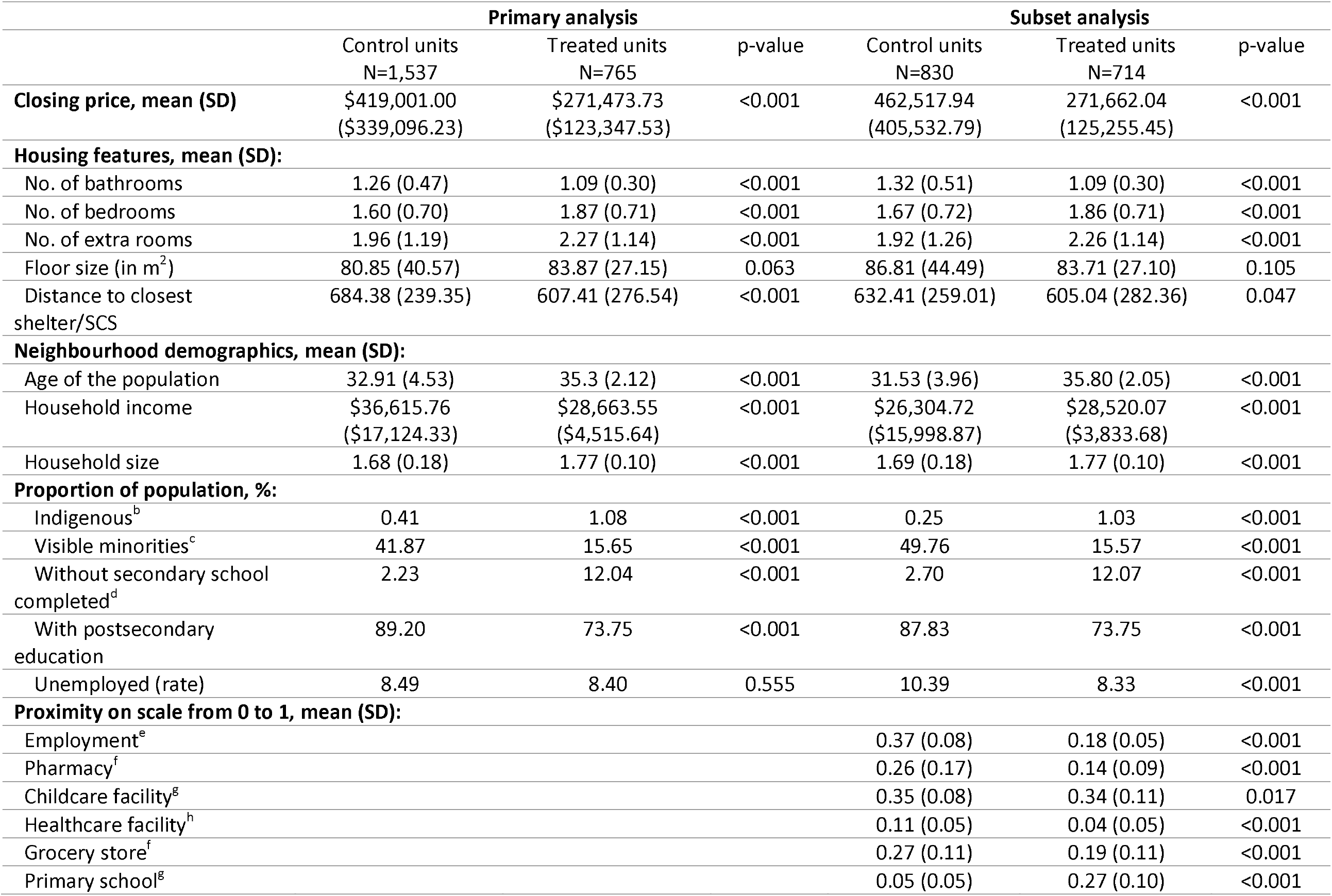
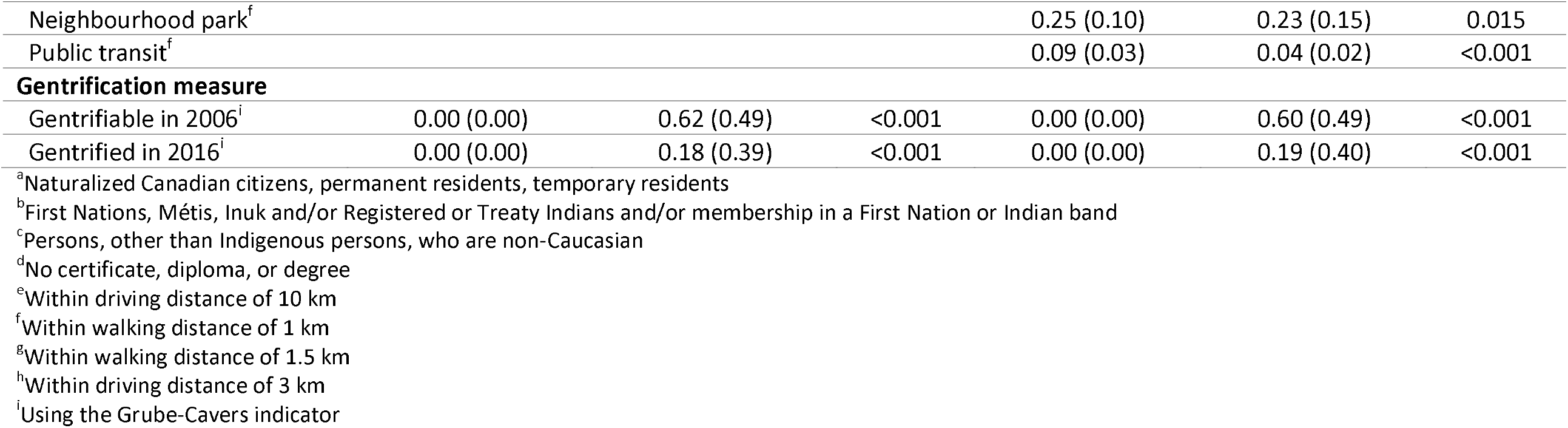
Comparison of house and neighborhood features of treated and control units *pre-intervention* (1 January 2014 – 31 July 2017)

|  | Primary analysis |  |  | Subset analysis |  |  |
| --- | --- | --- | --- | --- | --- | --- |
|  | Control units<br>N=1,537 | Treated units<br>N=765 | p-value | Control units<br>N=830 | Treated units<br>N=714 | p-value |
| **Closing price, mean (SD)** | \$419,001.00<br>(\$339,096.23) | \$271,473.73<br>(\$123,347.53) | <0.001 | 462,517.94<br>(405,532.79) | 271,662.04<br>(125,255.45) | <0.001 |
| **Housing features, mean (SD):** |  |  |  |  |  |  |
| No. of bathrooms | 1.26 (0.47) | 1.09 (0.30) | <0.001 | 1.32 (0.51) | 1.09 (0.30) | <0.001 |
| No. of bedrooms | 1.60 (0.70) | 1.87 (0.71) | <0.001 | 1.67 (0.72) | 1.86 (0.71) | <0.001 |
| No. of extra rooms | 1.96 (1.19) | 2.27 (1.14) | <0.001 | 1.92 (1.26) | 2.26 (1.14) | <0.001 |
| Floor size (in m<sup>2</sup>) | 80.85 (40.57) | 83.87 (27.15) | 0.063 | 86.81 (44.49) | 83.71 (27.10) | 0.105 |
| Distance to closest shelter/SCS | 684.38 (239.35) | 607.41 (276.54) | <0.001 | 632.41 (259.01) | 605.04 (282.36) | 0.047 |
| **Neighbourhood demographics, mean (SD):** |  |  |  |  |  |  |
| Age of the population | 32.91 (4.53) | 35.3 (2.12) | <0.001 | 31.53 (3.96) | 35.80 (2.05) | <0.001 |
| Household income | \$36,615.76<br>(\$17,124.33) | \$28,663.55<br>(\$4,515.64) | <0.001 | \$26,304.72<br>(\$15,998.87) | \$28,520.07<br>(\$3,833.68) | <0.001 |
| Household size | 1.68 (0.18) | 1.77 (0.10) | <0.001 | 1.69 (0.18) | 1.77 (0.10) | <0.001 |
| **Proportion of population, %:** |  |  |  |  |  |  |
| Indigenous<sup>b</sup> | 0.41 | 1.08 | <0.001 | 0.25 | 1.03 | <0.001 |
| Visible minorities<sup>c</sup> | 41.87 | 15.65 | <0.001 | 49.76 | 15.57 | <0.001 |
| Without secondary school completed<sup>d</sup> | 2.23 | 12.04 | <0.001 | 2.70 | 12.07 | <0.001 |
| With postsecondary education | 89.20 | 73.75 | <0.001 | 87.83 | 73.75 | <0.001 |
| Unemployed (rate) | 8.49 | 8.40 | 0.555 | 10.39 | 8.33 | <0.001 |
| **Proximity on scale from 0 to 1, mean (SD):** |  |  |  |  |  |  |
| Employment<sup>e</sup> |  |  |  | 0.37 (0.08) | 0.18 (0.05) | <0.001 |
| Pharmacy<sup>f</sup> |  |  |  | 0.26 (0.17) | 0.14 (0.09) | <0.001 |
| Childcare facility<sup>g</sup> |  |  |  | 0.35 (0.08) | 0.34 (0.11) | 0.017 |
| Healthcare facility<sup>h</sup> |  |  |  | 0.11 (0.05) | 0.04 (0.05) | <0.001 |
| Grocery store<sup>f</sup> |  |  |  | 0.27 (0.11) | 0.19 (0.11) | <0.001 |
| Primary school<sup>g</sup> |  |  |  | 0.05 (0.05) | 0.27 (0.10) | <0.001 |
| Neighbourhood park<sup>f</sup> |  |  |  | 0.25 (0.10) | 0.23 (0.15) | 0.015 |
| Public transit<sup>f</sup> |  |  |  | 0.09 (0.03) | 0.04 (0.02) | <0.001 |
| **Gentrification measure** |  |  |  |  |  |  |
| Gentrifiable in 2006<sup>i</sup> | 0.00 (0.00) | 0.62 (0.49) | <0.001 | 0.00 (0.00) | 0.60 (0.49) | <0.001 |
| Gentrified in 2016<sup>i</sup> | 0.00 (0.00) | 0.18 (0.39) | <0.001 | 0.00 (0.00) | 0.19 (0.40) | <0.001 |

<sup>a</sup>Naturalized Canadian citizens, permanent residents, temporary residents
<sup>b</sup>First Nations, Métis, Inuk and/or Registered or Treaty Indians and/or membership in a First Nation or Indian band
<sup>c</sup>Persons, other than Indigenous persons, who are non-Caucasian
<sup>d</sup>No certificate, diploma, or degree
<sup>e</sup>Within driving distance of 10 km
<sup>f</sup>Within walking distance of 1 km
<sup>g</sup>Within walking distance of 1.5 km
<sup>h</sup>Within driving distance of 3 km
<sup>i</sup>Using the Grube-Cavers indicator

In both the main and subset analyses, control houses had more bathrooms compared to treated units, while treated units had more rooms. Control neighbourhoods were younger with smaller household sizes, more ethnically diverse (larger proportion of population identified as visible minority), and more educated (larger proportion had post-secondary education). Somewhat paradoxically, control neighbourhoods also had a larger proportion of the population unemployed. Meanwhile, treated neighbourhoods had more indigenous residents. Control neighbourhoods had better proximity to employment opportunities, pharmacies, healthcare facilities, grocery stores, and public transit; while treatment neighbourhoods had better proximity to primary schools. Finally, treatment neighbourhoods were more likely to be identified as gentrifiable in 2006 and to have recently experienced gentrification.

After controlling for house and neighbourhood attributes (census tract data, and gentrification measure), and correcting for spatio-temporal correlation between records (Model 1), we found prior to the implementation of the local SCS, the closing price of treated units was 26.8% lower (95% CI: 23.5%, 29.9%) than control units’, with a month to month decrease of 0.2% (95% CI: -0.3%, 0.0%) compared to a 0.2% increase (95% CI: 0.1%, 0.2%) increase in the control units (Table 2). Following SCS implementation, there was an immediate (level) difference in treated compared with control unit prices, with a drop of 5.2% (95% CI: 1.4%, 8.8%) in sales price. However, treated units enjoyed a 0.6% increase (95% CI: 0.4%, 0.7%) in monthly price trends relative to controls (0.3%; 95% CI: 0.2%, 0.4%). When we controlled for proximity scores (Model 3), we found the level effect post-implementation disappeared (−3.3%, 95% CI: -0.7%, 1.1%), but monthly trend gains persisted (0.9%, 95% CI: 0.7%, 1.0%).

**Table 2:** Results of variations of controlled interrupted time series adjusting for housing and neighborhood attributes, without and with proximity measures (models 1 and 2 vs. 3 and 4, respectively), and restricted to pre-March 2020 sales (models 2 and 4).

|  | **Model 1<sup>a</sup>**<br>(95% CI) | **Model 2<sup>b</sup>**<br>(95% CI) | **Model 3<sup>c</sup>**<br>(95% CI) | **Model 4<sup>d</sup>**<br>(95% CI) |
| --- | --- | --- | --- | --- |
| Intercept ($\beta_0$) | **\$44,602**<br>**(\$34,419, \$57,797)** | **\$44,238**<br>**(\$32,288, \$60,611)** | **\$93,330**<br>**(\$64,537, \$134,968)** | **\$70,231**<br>**(\$44,636, \$110,504)** |
| Time ($\beta_1$) | **1.002**<br>**(1.001, 1.002)** | 1.001<br>(1.000, 1.002) | **1.003**<br>**(1.002, 1.004)** | **1.002**<br>**(1.001, 1.004)** |
| Group ($\beta_2$) | **0.732**<br>**(0.701, 0.765)** | **0.761**<br>**(0.725, 0.798)** | **0.722**<br>**(0.680, 0.766)** | **0.716**<br>**(0.668, 0.766)** |
| Group*Time ($\beta_3$) | **0.998**<br>**(0.997, 1.000)** | 0.999<br>(0.998, 1.000) | **0.997**<br>**(0.995, 0.998)** | **0.997**<br>**(0.996, 0.999)** |
| Level ($\beta_4$) | **1.077**<br>**(1.054, 1.101)** | 1.001<br>(0.976, 1.027) | **1.045**<br>**(1.015, 1.076)** | 0.986<br>(0.953, 1.021) |
| Trend ($\beta_5$) | **1.003**<br>**(1.002, 1.004)** | **1.007**<br>**(1.005, 1.008)** | 1.000<br>(0.999, 1.001) | **1.003**<br>**(1.001, 1.004)** |
| SCS level ($\beta_6$) | **0.948**<br>**(0.912, 0.986)** | 1.000<br>(0.955, 1.047) | 0.967<br>(0.925, 1.011) | 0.997<br>(0.945, 1.051) |
| SCS trend ($\beta_7$) | **1.006**<br>**(1.004, 1.007)** | **1.003**<br>**(1.000, 1.005)** | **1.009**<br>**(1.007, 1.010)** | **1.008**<br>**(1.005, 1.010)** |
| Pseudo-$R^2$ | 0.837 | 0.842 | 0.851 | 0.857 |
| $n$ | 6,903 | 4,801 | 4,815 | 3,313 |

<sup>a</sup>Controlling for housing and neighbourhood attributes with spatio-temporal price lag, excluding proximity scores
<sup>b</sup>Controlling for housing and neighbourhood attributes with spatio-temporal price lag; restricted to sales pre-March 2020, excluding proximity scores
<sup>c</sup>Controlling for housing and neighbourhood attributes with spatio-temporal price lag, including proximity scores
<sup>d</sup>Controlling for housing and neighbourhood attributes with spatio-temporal price lag; restricted to sales pre-March 2020, including proximity scores
**bold** indicates statistical significance

In other words, among treated sales we observed an absolute increase of $37,931.86 (95% CI: $12,223.35, $138,088.50) by December 2021 (relative change: 53.3%, 95% CI: 33.4%, 76.3%) compared with the counterfactual scenario (i.e., had the SCS not been implemented). For sales retained in the subset analysis that controlled for proximity to amenities, treated sales were $12,224.11 higher (95% CI: $3,849.57, $30,796.24) than expected by December 2021 (relative change: 30.1%, 95% CI: 15.4%, 46.7%).

## Discussion

Between January 2016 and June 2022, 32,632 Canadians died of opioid toxicity (Special Advisory Committee on the Epidemic of Opioid Overdoses, 2022). Despite the ongoing urgency of the overdose crisis and plethora of studies demonstrating SCSs’ effectiveness in mitigating drug use related morbidity and mortality, communities continue to resist their implementation. Our study directly examined one ongoing aspect of opponents’ arguments against SCSs – that their presence jeopardizes residential real estate value.

Our results indicate that neighbourhoods where SCSs were implemented were qualitatively different from control units’ neighbourhoods. Specifically, treated sales were in neighbourhoods where households were larger; and the population was older, less ethnically diverse, and with fewer adults who had completed secondary school. Treated neighbourhoods also had lower proximity to employment opportunities, pharmacies, healthcare facilities, and public transit, and were identified as more likely to undergo gentrification. Together, these features suggest treated neighbourhoods were economically more depressed compared with control neighbourhoods, prior to SCS implementation.

Our analysis found an immediate negative effect of SCSs on residential real estate prices. However, closing prices of properties within 1,000m of SCSs rose faster than those in control neighbourhoods, indicating community wide benefits of SCS implementation (Table 2, Figure 2). These results contradict findings recently reported by Liang and Alexeev who found a 5 – 7% reduction in residential real estate closing prices in Victoria, Australia (Liang & Alexeev, 2023). However, their study significantly differed from ours in several ways. The team used a difference-in-differences study design that did not effectively account for the underlying temporal trends between treated and control units, considered treated sales within 800m of their SCS and control sales 800 – 2,000m beyond this radius, did not account for many critical unit- and neighbourhood-level attributes that would otherwise explain differences in closing sales prices, and neglected to control for spatio-temporal correlation between sales – potentially inflating estimated effects.

**Figure 2:**
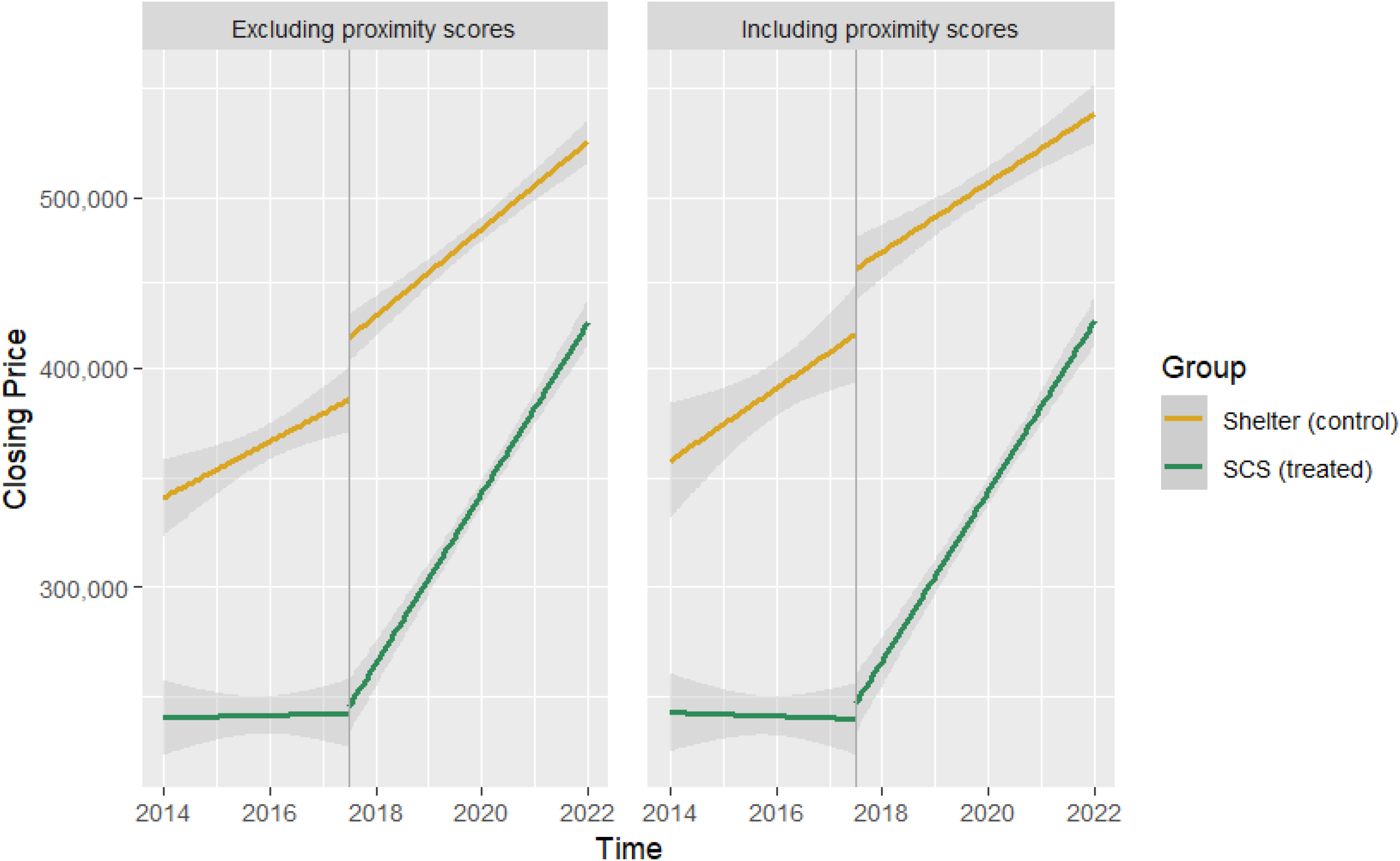
Comparison of price in treated vs. control sales pre- vs. post-intervention implementation controlling for house and neighborhood amenities, with and without proximity scores in models

Prior research demonstrates the effects of (dis)amenities are distance dependent (e.g., housing prices within the first 400m of a commuter train station decline, while prices immediately outside this radius (e.g., 400 – 800m) but still within ‘walking distance’ of the station, increase)(Dubé et al., 2017). Marshall et al.’s seminal study observed that over 70% of clients of Vancouver’s Insite were within 4 blocks of the supervised consumption site (Marshall BDL et al., 2011), and more recent evaluations of SCSs have observed impacts on health service use within 500m of sites (The Evaluation of Overdose Prevention Sites Working Group & Lori Wagar, 2018). These studies suggest that the distances used to distinguish treated and control units in the Liang and Alexeev paper may be too large; effectively comparing very disparate housing markets without appropriately accounting for these differences. Meanwhile, the magnitudes and directions of externalities’ effects on housing prices depend on the socio-economic status of the neighbourhood (e.g., the commercial activity generated by a commuter train station may positively impact a low-income neighbourhood but have little to no positive impact (or even negative impact) on housing prices in an affluent part of town) (Forouhar & Hasankhani, 2018). All three of Montreal’s SCSs were implemented in urban neighbourhoods very close to the city’s downtown core. Although our control units were also urban and within 1,000m of men’s homeless shelters, treated units were effectively in more economically depressed neighbourhoods. Conversely, Victoria’s SCS was implemented in a peri-urban neighbourhood with insufficient information to determine socio-economic status of treated sales’ neighbourhoods compared with control sales. Differences in the location of the SCSs may therefore also explain why we observed a positive effect of SCSs on residential real-estate prices post-implementation and Liang and Alexeev did not.

Our study had several strengths. We used advancements in econometrics to account for traditionally neglected spatio-temporal correlation for a more nuanced examination of consumers’ revealed preferences. Failing to account for the two-dimensional correlation structure of our data could bias the results and overestimate the effect of the intervention. We also examined our records to understand the magnitude and potential effect of house flipping – and observed limited impact of this phenomenon in our sales records. By focusing on a large city with a dispersed population of PWUD and the effects of SCSs across multiple neighbourhoods, we were able to account for the potential honey-pot effect and reproducibility of our results, respectively. Further, by selecting a city that was not experiencing a frenzied housing market during the observation period, we reduced the potential for a hot housing market to obscure the effects of the intervention. Finally, using over three years of pre- and post-implementation data sufficiently powered our study to observe very small but plausibly significant effects of SCSs on real estate prices given the compounding effects of trends in housing prices over time.

Limitations of our study included the exclusion of approximately 4.9% (n =566) of sales records owing to data quality challenges in the initial data cleaning. However, as part of data clean up and preparation efforts, we noted no observable differences in the quality of data between treated and control units (i.e., differential exclusion). During analysis, missing data led to an additional 16.7% (n=1,413) of records being excluded, with a further 25.1% (n=2,088) records excluded in the subset that includes proximity scores. Further, despite SCSs operating in almost every province in Canada, we were unable to secure sales records from other cities. This may restrict the generalizability of our findings. Finally, although we note a positive trend in monthly housing price growth in treated neighbourhoods post-implementation we can only postulate on why. Elsewhere, SCSs have been shown to improve local communities’ physical environments via reductions in public drug use and drug-related litter.(Freeman et al., 2005; Wood et al., 2004)

Our study provides evidence that the implementation of SCSs can have a positive impact on local residential real estate prices, with prices of houses sold in neighbourhoods with an SCS rising faster than those in control neighbourhoods. This phenomenon may reflect improvements in the physical environment and overall quality of life in the area. However, more research is needed to understand the mechanisms behind this effect, whether it is unique to the Montreal context, and the long-term sustainability of this trend. Nevertheless, our results provide a valuable contribution to the current debate surrounding SCSs and their impacts on local communities.

## Data Availability

Due to commercial restrictions, supporting data is not available.

