## Supplemental Tables for "Evaluating the effects of supervised consumption sites on housing prices in Montreal, Canada using controlled interrupted time series and hedonic price models"

**Supplementary Material**

Table 1: Controlled interrupted time series results (excluding proximity scores)

|  | **Model 1^a^**  **(95% CI)** | **Model 2^b^**  **(95% CI)** | **Model 3^c^**  **(95% CI)** | **Model 4^d^**  **(95% CI)** |
| --- | --- | --- | --- | --- |
| Intercept ($\beta_{0})$ | **$383,259**  **($369,431, $397,605)** | **$42,203**  **($32,514, $54,780)** | **$44,602**  **($34,419, $57,797)** | **$44,238**  **($32,288, $60,611)** |
| Time ($\beta_{1})$ | **1.003**  **(1.001, 1.005)** | **1.002**  **(1.002, 1.003)** | **1.002**  **(1.001, 1.002)** | 1.001  (1.000, 1.002) |
| Group ($\beta_{2})$ | **0.663**  **(0.622, 0.707)** | **0.714**  **(0.683, 0.745)** | **0.732**  **(0.701, 0.765)** | **0.761**  **(0.725, 0.798)** |
| Group*Time ($\beta_{3})$ | **0.997**  **(0.994, 1.000)** | **0.998**  **(0.996, 0.999)** | **0.998**  **(0.997, 1.000)** | 0.999  (0.998, 1.000) |
| Level ($\beta_{4})$ | **1.083**  **(1.033, 1.136)** | **1.076**  **(1.053, 1.100)** | **1.077**  **(1.054, 1.101)** | 1.001  (0.976, 1.027) |
| Trend ($\beta_{5})$ | 1.002  (1.000, 1.004) | **1.003**  **(1.002, 1.004)** | **1.003**  **(1.002, 1.004)** | **1.007**  **(1.005, 1.008)** |
| SCS level ($\beta_{6})$ | 0.927  (0.852, 1.009) | **0.946**  **(0.910, 0.984)** | **0.948**  **(0.912, 0.986)** | 1.000  (0.955, 1.047) |
| SCS trend ($\beta_{7})$ | **1.008**  **(1.004, 1.011)** | **1.006**  **(1.004, 1.007)** | **1.006**  **(1.004, 1.007)** | **1.003**  **(1.000, 1.005)** |
| **Housing features:** |  |  |  |  |
| No. of bathrooms |  | **1.214**  **(1.196, 1.233)** | **1.211**  **(1.193, 1.229)** | **1.206**  **(1.184, 1.228)** |
| No. of bedrooms |  | **1.097**  **(1.087, 1.107)** | **1.095**  **(1.085, 1.104)** | **1.095**  **(1.083, 1.107)** |
| No. of extra rooms |  | **1.008**  **(1.005, 1.012)** | **1.008**  **(1.005, 1.012)** | **1.008**  **(1.004, 1.013)** |
| Floor size (in m2) |  | **1.007**  **(1.006, 1.007)** | **1.007**  **(1.006, 1.007)** | **1.007**  **(1.007, 1.007)** |
| Distance to closest shelter/SCS |  | **1.000**  **(1.000, 1.000)** | **1.000**  **(1.000, 1.000)** | **1.000**  **(1.000, 1.000)** |
| **Neighbourhood demographics:** |  |  |  |  |
| Age of the population |  | **1.003**  **(1.002, 1.004)** | **1.005**  **(1.004, 1.007)** | **1.006**  **(1.004, 1.008)** |
| Household income |  | **1.000**  **(1.000, 1.000)** | **1.000**  **(1.000, 1.000)** | **1.000**  **(1.000, 1.000)** |
| Household size |  | **1.191**  **(1.131, 1.253)** | **1.213**  **(1.153, 1.277)** | **1.181**  **(1.111, 1.255)** |
| **Proportion of population, %:** |  |  |  |  |
| Visible minorities^e^ |  | **0.998**  **(0.996, 0.999)** | **0.997**  **(0.996, 0.999)** | **0.997**  **(0.996, 0.999)** |
| Without secondary school completed^f^ |  | **1.007**  **(1.003, 1.012)** | 1.003  (0.999, 1.008) | 1.003  (0.997, 1.008) |
| With postsecondary education |  | **1.016**  **(1.013, 1.018)** | **1.013**  **(1.011, 1.016)** | **1.013**  **(1.009, 1.016)** |
| Unemployed (rate) |  | **0.977**  **(0.974, 0.980)** | **0.978**  **(0.975, 0.980)** | **0.978**  **(0.974, 0.981)** |
| **Gentrification index** |  |  |  |  |
| Gentrifiable in 2006^k^ |  | **0.911**  **(0.891, 0.931)** | **0.914**  **(0.895, 0.94)** | **0.909**  **(0.885, 0.933)** |
| Gentrified in 2016^k^ |  | 0.993  (0.970, 1.017) | 0.998  (0.975, 1.022) | 1.001  (0.972, 1.030) |
| Pseudo-R2 | 0.234 | 0.835 | 0.837 | 0.842 |
| n | 6,903 | 6,903 | 6,903 | 4,801 |

^a^Crude model

^b^Controlling for housing and neighbourhood attributes

^c^Controlling for housing and neighbourhood attributes with spatio-temporal price lag

^d^Controlling for housing and neighbourhood attributes with spatio-temporal price lag; restricted to sales pre-March 2020

^e^ Persons who are non-Caucasian including Indigenous persons (First Nations, Métis, Inuk and/or Registered or Treaty Indians and/or membership in a First Nation or Indian band)

^f^No certificate, diploma, or degree

^g^Within driving distance of 10 km

^h^Within walking distance of 1 km

^i^Within walking distance of 1.5 km

^j^Within driving distance of 3 km

^k^Using the Grube-Cavers indicator

**bold** indicates statistical significance

Table 2: Controlled interrupted time series results (including proximity scores)

|  | **Model 1^a^**  **(95% CI)** | **Model 2^b^**  **(95% CI)** | **Model 3^c^**  **(95% CI)** | **Model 4^d^**  **(95% CI)** |
| --- | --- | --- | --- | --- |
| Intercept ($\beta_{0})$ | **$417,473**  **($396,879, $439,137)** | **$92,695**  **($63,925, $134,414)** | **$93,330**  **($64,537, $134,968)** | **$70,231**  **($44,636, $110,504)** |
| Time ($\beta_{1})$ | **1.004**  **(1.002, 1.006)** | **1.004**  **(1.003, 1.005)** | **1.003**  **(1.002, 1.004)** | **1.002**  **(1.001, 1.004)** |
| Group ($\beta_{2})$ | **0.605**  **(0.561, 0.652)** | **0.718**  **(0.676, 0.763)** | **0.722**  **(0.680, 0.766)** | **0.716**  **(0.668, 0.766)** |
| Group*Time ($\beta_{3})$ | **0.996**  **(0.992, 0.999)** | **0.996**  **(0.995, 0.998)** | **0.997**  **(0.995, 0.998)** | **0.997**  **(0.996, 0.999)** |
| Level ($\beta_{4})$ | **1.089**  **(1.022, 1.161)** | **1.049**  **(1.018, 1.080)** | **1.045**  **(1.015, 1.076)** | 0.986  (0.953, 1.021) |
| Trend ($\beta_{5})$ | 1.000  (0.997, 1.002) | 1.001  (1.000, 1.002) | 1.000  (0.999, 1.001) | **1.003**  **(1.001, 1.004)** |
| SCS level ($\beta_{6})$ | 0.932  (0.845, 1.028) | 0.959  (0.917, 1.003) | 0.967  (0.925, 1.011) | 0.997  (0.945, 1.051) |
| SCS trend ($\beta_{7})$ | **1.010**  **(1.006, 1.014)** | **1.008**  **(1.006, 1.010)** | **1.009**  **(1.007, 1.010)** | **1.008**  **(1.005, 1.010)** |
| **Housing features:** |  |  |  |  |
| No. of bathrooms |  | **1.194**  **(1.173, 1.216)** | **1.188**  **(1.167, 1.210)** | **1.190**  **(1.164, 1.216)** |
| No. of bedrooms |  | **1.090**  **(1.079, 1.102)** | **1.089**  **(1.078, 1.100)** | **1.090**  **(1.076, 1.104)** |
| No. of extra rooms |  | **1.008**  **(1.003, 1.012)** | **1.008**  **(1.003, 1.012)** | **1.006**  **(1.000, 1.011)** |
| Floor size (in m2) |  | **1.007**  **(1.006, 1.007)** | **1.007**  **(1.006, 1.007)** | **1.007**  **(1.006, 1.007)** |
| Distance to closest shelter/SCS |  | **1.000**  **(1.000, 1.000)** | **1.000**  **(1.000, 1.000)** | **1.000**  **(1.000, 1.000)** |
| **Neighbourhood demographics:** |  |  |  |  |
| Age of the population |  | 0.999  (0.997, 1.002) | 1.000  (0.998, 1.003) | 1.000  (0.997, 1.003) |
| Household income |  | **1.000**  **(1.000, 1.000)** | **1.000**  **(1.000, 1.000)** | **1.000**  **(1.000, 1.000)** |
| Household size |  | **1.276**  **(1.188, 1.371)** | **1.324**  **(1.232, 1.423)** | **1.387**  **(1.273, 1.512)** |
| **Proportion of population, %:** |  |  |  |  |
| Visible minorities^e^ |  | **0.994**  **(0.992, 0.995)** | **0.994**  **(0.992, 0.995)** | **0.993**  **(0.991, 0.995)** |
| Without secondary school completed^f^ |  | 1.001  (0.996, 1.006) | 0.999  (0.993, 1.004) | 1.002  (0.996, 1.009) |
| With postsecondary education |  | **1.004**  **(1.000, 1.008)** | 1.003  (0.999, 1.007) | **1.005**  **(1.001, 1.010)** |
| Unemployed (rate) |  | **0.972**  **(0.968, 0.976)** | **0.972**  **(0.968, 0.976)** | **0.971**  **(0.966, 0.975)** |
| **Proximity to:** |  |  |  |  |
| Employment^g^ |  | **4.795**  **(3.658, 6.285)** | **4.090**  **(3.118, 5.364)** | **3.543**  **(2.562, 4.900)** |
| Pharmacy^h^ |  | **0.645**  **(0.572, 0.728)** | **0.665**  **(0.590, 0.750)** | **0.634**  **(0.549, 0.734)** |
| Childcare facility^i^ |  | 1.025  (0.918, 1.144) | 0.989  (0.887, 1.104) | 0.906  (0.792, 1.037) |
| Healthcare facility^j^ |  | **1.795**  **(1.191, 2.707)** | **2.182**  **(1.448, 3.288)** | **2.634**  **(1.608, 4.315)** |
| Grocery store^h^ |  | **1.144**  **(1.049, 1.248)** | **1.190**  **(1.091, 1.298)** | **1.348**  **(1.214, 1.497)** |
| Primary school^i^ |  | 0.983  (0.892, 1.095) | 1.034  (0.928, 1.153) | 1.088  (0.951, 1.245) |
| Neighbourhood park^h^ |  | 0.947  (0.890, 1.008) | **0.936**  **(0.880, 0.996)** | 0.950  (0.880, 1.026) |
| Public transit^h^ |  | 0.847  (0.593, 1.210) | 0.733  (0.514, 1.046) | 0.963  (0.621, 1.492) |
| **Gentrification index** |  |  |  |  |
| Gentrifiable in 2006^k^ |  | **0.923**  **(0.900, 0.947)** | **0.926**  **(0.903, 0.949)** | **0.921**  **(0.893, 0.950)** |
| Gentrified in 2016^k^ |  | 0.976  (0.951, 1.002) | 0.978  (0.953, 1.004) | 0.985  (0.954, 1.017) |
| Pseudo-R2 | 0.277 | 0.849 | 0.851 | 0.857 |
| n | 4,815 | 4,815 | 4,815 | 3,313 |

^a^Crude model

^b^Controlling for housing and neighbourhood attributes

^c^Controlling for housing and neighbourhood attributes with spatio-temporal price lag

^d^Controlling for housing and neighbourhood attributes with spatio-temporal price lag; restricted to sales pre-March 2020

^e^Persons who are non-Caucasian including Indigenous persons (First Nations, Métis, Inuk and/or Registered or Treaty Indians and/or membership in a First Nation or Indian band)

^f^No certificate, diploma, or degree

^g^Within driving distance of 10 km

^h^Within walking distance of 1 km

^i^Within walking distance of 1.5 km

^j^Within driving distance of 3 km

^k^Using the Grube-Cavers indicator

**bold** indicates statistical significance
